# The Role of Parenting in Mitigating Epigenetic Cardiometabolic Risk in a Sample of Predominantly Latino Preschoolers

**DOI:** 10.64898/2026.07.17.26358350

**Authors:** Adamari López, Sarah M. Merrill, Anne K. Bozack, Andres Cardenas, Jonathan S. Comer, Daniel M. Bagner, April Highlander, Justin Parent

## Abstract

**Background:** Childhood obesity is a prevalent public health concern, particularly among children from marginalized backgrounds. Epigenomic mechanisms, including DNA methylation (DNAm), offer insight into the biological embedding of metabolic risk, yet protective family-level factors remain understudied. This study examined whether participation in a positive parenting intervention was associated with reduced epigenetic cardiometabolic risk among preschool-aged children.

**Methods:** Participants were 74 children (*n* = 35 intervention; *n* = 39 services-as-usual; mean age = 36 months). Using a secondary analysis of a randomized controlled trial, DNAm-derived body mass index (BMI) and anthropometric BMI were assessed at baseline and 12-month follow-up, and parenting practices were observed post-treatment.

**Results:** Children in the intervention group demonstrated significantly lower DNAm BMI at 12 months relative to controls, adjusting for child sex, race and/or ethnicity, and anthropometric BMI. No treatment effect was observed for anthropometric BMI, and DNAm BMI was not associated with anthropometric BMI at 12 months. Although parenting practices improved, they did not mediate intervention effects on DNAm-derived BMI.

**Conclusions:** Parenting interventions may influence biological pathways related to cardiometabolic risk, even before changes in anthropometric outcomes, underscoring the potential of family-centered approaches to promote early metabolic health.

**Impact:**

- Participation in a positive parenting intervention was associated with a lower epigenetic biomarker of BMI in preschool-aged children.
- Findings highlight a disconnect between biological markers and traditional anthropometric measures.
- Results underscore potential for early family-centered interventions to shape biological pathways associated with long-term health, ultimately supporting prevention strategies before clinical risk is observable.

## Introduction

Childhood obesity remains a significant public health issue in the United States, with national data indicating that approximately 1 in 5 children and adolescents (ages 2-19) meet criteria for obesity via measurement of body-mass-index (BMI), translating to nearly 14.7 million youth.^1^ Within preschool-aged children specifically, 12.7% of U.S. children between 2 and 5 years old^2^ meet criteria for obesity. Prevalence rates remain disproportionately high among children from marginalized racial/ethnic groups and socioeconomically disadvantaged backgrounds, underscoring persistent disparities in risk and outcomes.^3,4^ Latino youth specifically are among the ethnic groups with the highest prevalence of elevated weight status or risk for obesity in the United States, with 45.5% of Latino children falling in the overweight range and 23.5% meeting criteria for obesity.^5,6^ Importantly, childhood obesity is a developmental precursor to adulthood obesity, heightened risk for difficulties with cardiovascular health across the lifespan,^7,8^ increased risk for other physiological health problems such as asthma, diabetes, liver disease, reproductive problems, and some cancers,^9^ and increased risk for psychological health concerns such as social isolation and depression.^10,11^ As Latino individuals make up almost one-fifth (19.5%) of the U.S. population and constitute the second largest ethnic group in the U.S. after the non-Latino White population, addressing these health disparities is a public health concern with implications across the lifespan.

Despite ample empirical support for measuring obesity and cardiometabolic risk via standard weight- and height-derived BMI (i.e., anthropometrically derived BMI), there have been critiques of its validity due to its lack of generalizability to minoritized populations.^12^ In these contexts, anthropometric BMI may overestimate cardiometabolic risk because it fails to account for structural and contextual influences such as socioeconomic status inequities that shape health beyond body size alone. Furthermore, anthropometric BMI clinical cut-offs and clinical interpretations have been largely developed based on White European samples, which may fail to adequately account for ancestry-related differences in body composition, adipose tissue distribution, and bone density.^13^ Although anthropometric BMI is often used as an indicator of obesity risk, it does not directly measure body composition, meaning that it cannot distinguish adipose and lean mass or capture underlying biological processes relevant to cardiometabolic health. Changes in anthropometric BMI and metabolic biomarkers, such as insulin resistance and lipid levels, typically become detectable only after obesity has progressed or the body has experienced considerable physiological dysregulation^14^. As such, current clinical tools may both be biased and insufficient for revealing early molecular vulnerability among racially and ethnically marginalized young children.

As an alternative to anthropometric BMI and current obesity categories, epigenetic obesity risk scores offer a health-centric approach to child cardiometabolic risk.^15^ Epigenetics provides a bridge between biology and environment, illustrating how experiences and contexts become biologically embedded to shape health trajectories.^16^ One key mechanism, DNA methylation (DNAm), often involves the addition of methyl groups at cytosine–phosphate–guanine (CpG) dinucleotides, thereby affecting gene expression or transcriptional activity but not the underlying DNA sequence. Because DNAm patterns are sensitive to environmental and psychosocial exposures, they offer a valuable framework for identifying both biological risk and resilience in relation to long-term health outcomes. Obesity and cardiometabolic risk are public health concerns that can be examined through a bioecological lens, which suggests that child development arises from continuous interactions between biological processes and layered environmental systems, including family, community, and sociocultural contexts.^17^ This perspective highlights the importance of considering not only individual-level behaviors but also the ecological conditions that shape risk. Recent large-scale work on Latino cardiovascular health further demonstrates how sociocultural, psychological, and contextual factors shape cardiometabolic outcomes, reinforcing the need for models that move beyond individual-level indicators such as anthropometric BMI.^18^ Epigenetic methods align with this framework by providing a modern approach to directly assess the molecular connection between ecological factors and child biology. Adopting this approach grants us the ability to examine individuals within their contexts, taking into consideration biological predisposition, as well as the strong influences of family and peers (e.g., family modeling of physical activity, food habits, sleep), community environments (e.g., parks, green space, public transport and food outlets), and society or public policy (e.g., government policies, food marketing, transport systems), instead of viewing individuals or genetic predisposition as sole actors in determining BMI or cardiometabolic risk.^19,20^

Initial research applying this bioecological lens demonstrates that adverse environmental exposures can become biologically embedded, influencing epigenetic markers linked to obesity risk. For instance, adverse childhood experiences (ACEs) have been associated not only with increased obesity in youth,^20,21^ but also changes in DNAm at sites related to anthropometric BMI.^22,23^ Similarly, adolescents from socioeconomically underserved and disadvantaged backgrounds demonstrate higher epigenetic BMI risk scores, suggesting that early environmental adversity may influence biological systems in ways that heighten cardiometabolic vulnerability.^24^ Importantly, Raffington and colleagues^24^ found that early-life adversity during infancy predicted epigenetic BMI scores in middle childhood, which was linked to a higher risk of excess weight in later adolescence. However, no research to our knowledge has examined epigenetic BMI scores in early childhood nor explored the role of environmental factors beyond SES, leaving critical gaps in understanding the early development of these epigenetic markers and health trajectories.

Building on this emerging evidence, the family environment, specifically the quality of parent-child interactions, represents a key proximal context through which broader environmental factors may become biologically embedded. Parent-child interactions can initiate a cascade of biological changes, beginning with changes in stress reactivity. Responsive and consistent parenting practices (e.g., warmth, praise, discipline) have been associated with healthier weight outcomes and cardiometabolic health.^25,26^ Conversely, harsh and inconsistent parenting practices such as parental demandingness, overprotection, and psychological control demonstrated associations with less desirable health outcomes (i.e., higher body weight)^27^. Further, lower levels of harsh and inconsistent parenting practices^28,29,29,30^ and higher levels of responsive and consistent parenting^30^ have been found to be associated with child epigenetic aging or stress-related DNAm risk scores (e.g., HPA axis, inflammation). These patterns may reflect how emotionally supportive and structured environments buffer children from chronic stress exposure and promote adaptive physiological regulation, reducing the biological embedding of obesity risk.

While observational studies suggest important associations between parenting and both cardiometabolic health and epigenetic outcomes, intervention research offers stronger evidence for causal pathways. For example, interventions that strengthen general parenting skills – such as programs emphasizing responsive caregiving and consistent limit-setting (e.g., Parent-Child Interaction Therapy [PCIT]) – have demonstrated promise in improving family functioning and mitigating child cardiometabolic risk.^31–33^ Furthermore, these programs and others that strengthen the parent-child relationship have been shown to positively influence child epigenetic aging or stress-related DNAm outcomes,^34–36^ as well as buffer the impact of adversity on these epigenetic outcomes,^37,38^ although no studies have examined epigenetic BMI outcomes. Therefore, parenting practices may represent a modifiable factor through which children’s biological susceptibility to obesity can be influenced across development.

## Current Study

Although prior research has established links between parenting and childhood obesity, stress and obesity, and parenting and child epigenetic outcomes, much less is known about how parenting relates to epigenetic patterns associated with resilience and healthy cardiometabolic functioning. No studies to our knowledge have examined how changes in parenting influence DNAm-based BMI risk scores, which may be particularly vital to study in early childhood when obesity risk trajectories are being established.^39^ Addressing this gap is especially urgent for those from marginalized backgrounds, such as Latino youth and those from economically under-resourced households, given that they experience disproportionally high rates of childhood obesity^40^ yet remain significantly underrepresented in epigenetic research.^41^ The current study aims to examine whether participation in an evidence-based parenting intervention is associated with lower DNAm-based BMI risk scores among predominantly Latino preschoolers from marginalized racial, ethnic, and socioeconomic backgrounds. Furthermore, the current study aims to investigate whether improvements in specific parenting behaviors (e.g., warmth, consistent discipline, and emotional support) might mediate the relation between group (i.e., iPCIT vs. RAU) and changes in metabolic risk. This study will represent one of the first examinations of whether a parenting intervention influences DNAm-based BMI risk scores in early childhood, with implications for developing prevention strategies that address the biological embedding of obesity risk in marginalized communities.

## Methods

### Participants

The current sample included *N* = 74 primary caregivers and their 3-year-old children. Participants were recruited from an ongoing RCT^42^ examining the impacts of a telehealth-based parenting intervention^43^ (Internet-delivered PCIT, or iPCIT) on child outcomes, including child epigenetic outcomes.^35,37^ For inclusion, children in the study exhibited elevated levels of disruptive behavior and developmental delay, a population at heightened risk for obesity^44^ and adverse health outcomes.^45^ Participants were recruited between early 2016 and mid 2019 from Early Intervention sites in the greater Miami-Dade area. Follow-up sessions were held through December 2020. Caregiver-child dyads were randomly assigned to the parenting intervention group or the control group. Inclusion criteria for the study consisted of parent-reported Child Behavior Checklist (CBCL) Externalizing Problems^46^ T-score greater than 60 and primary caregivers speaking English or Spanish. Exclusion criteria consisted of: children receiving medication for behavior problems, child or caregiver deafness or blindness, severe child social communication deficits measured via the Social Responsiveness Scale (SRS), second edition^47^ (T score > 75), and primary caregiver standard score of less than 4 on the vocabulary subtest of the Wechsler Abbreviated Scale of Intelligence (WASI-II).^48^

### Procedure

Upon beginning the study, caregiver-child dyads were randomly assigned to either the iPCIT group (See below for more details) for 20 weeks or the referrals as usual (RAU) control group. The study’s major assessments were conducted in the family’s home at four distinct time points—baseline, week 20 (i.e., post), 6-month, and 12-month post-treatment follow-ups. The present study utilized data from the baseline and 12-month follow-up assessments as the 12-month assessment represented the primary long-term follow-up time point and provided the most complete data for the variable included in the current analyses. Families received $100 for each major assessment and an electronic tablet at study completion. Informed consent for the DNA sub-study of the RCT was obtained at either baseline, post-treatment, or follow-up home visits, depending on families’ progress in the intervention at the time of institutional review board approval for the DNA sub-study. All participants completed the treatment phase before the COVID-19 pandemic.

### Parenting Intervention

Parent-Child Interaction Therapy (PCIT) is an evidence-based parenting program with strong empirical support for enhancing responsive and consistent parenting practices across diverse populations.^49^ Internet-based Parent-Child Interaction Therapy (iPCIT)^43^ expands accessibility by using HIPAA-compliant videoconferencing, enabling therapists to remotely provide live coaching and feedback on caregiver-child interactions in their own homes. Congruent with clinic-based PCIT, iPCIT progresses through two phases of treatment: child-directed interactions and parent-directed interactions. During child-directed interactions, caregivers learn to follow their child’s lead in play by using praise, warmth, and child-focused attention strategies, while avoiding commands, criticisms, and questions. In the parent-directed interaction phase, caregivers learn to use effective commands, limit setting, and consistency in enforcing consequences to increase child compliance. iPCIT has demonstrated significant, sustained improvements in child behavior problems and caregiving behaviors among families from historically underserved communities.^42^

## Measures

### Independent Variables

The primary independent variable in this study is treatment group assignment. Families were randomly enrolled in one of two conditions: iPCIT or RAU. Participants in the iPCIT group received the full two-phase intervention for up to 20 weeks (average sessions completed: *M* = 9.44), delivered via secure videoconferencing, as described above. In contrast, families assigned to the RAU group were provided with standard community referrals without additional intervention from the study team. Treatment group assignment allows for examination of the effects of receiving iPCIT on child and parent outcomes compared with typical service pathways.

#### Mediators

Parenting behaviors were assessed using observational methods. As part of the RCT assessment protocol, dyads participated in a 5-minute child-led play task at baseline and post-treatment, which was coded using the Dyadic Parent–Child Interaction Coding System, Fourth Edition (DPICS-IV).^50^ The DPICS-IV is a standardized observational instrument designed to evaluate the quality of parent–child interactions, with established reliability and validity in assessing parenting skills in diverse families. Responsive and consistent parenting was assessed by “Do” skills during child-led play, which include labeled praises (e.g., “Great job cleaning up!”), behavior descriptions (e.g., “I noticed you shared your building blocks with your friend.”, and reflections (e.g., “I see that you’re sad that play time is over.”, whereas harsh and inconsistent parenting was assessed by “don’t” skills, including questions, commands, and criticisms. At each major assessment, the proportion of caregiver “do” and “don’t” skills during a 5-minute child-led play task was coded to reflect consistent versus inconsistent parenting behaviors (mean κ = 0.83 across codes for total sample).

### Dependent Variables

Salivary samples were collected from children during all home visits, with baseline and 12-month follow-up visits of particular interest to the current study, using Oragene kits (ORG-575) for assisted collection (DNA Genotek). DNA methylation (DNAm) was measured at the University of Minnesota Genomics Center. Following bisulfate conversion, DNAm was measured using the Infinium HumanMethylationEPIC Bead Chip array. Assayed data were processed with R version 4.1.1 (R Project for Statistical Computing) with packages minfi^51^ and ewastools^52^ as previously described.^35^ Normalization was performed with funnorm with noob for background and color adjustment.^53^ Buccal epithelial cell (BEC) proportion was estimated using EpiDISH with robust partial correlation.^54^ Child DNAm-derived BMI was assessed using the McCartney et al.^55^ BMI risk score. These risk scores were developed using LASSO penalized regression to find CpG sites whose DNAm levels were strongly associated with measured BMI in adults. These DNAm BMI scores capture molecular signatures of adiposity and metabolic regulation and correlate strongly with cardiometabolic markers (e.g., insulin resistance, inflammation). Further, Raffington et al^24^ found that DNAm-derived BMI risk scores correlate with children’s BMI and socioeconomic status, indicating sensitivity to potential environmental stressors.

Child anthropometric BMI was measured at each in-home data collection visit. During in-home visits, trained research staff obtained child height and weight using standardized anthropometric procedures. A standard scale was used to measure child weight, and a standard tape measure was used to measure child height. Anthropometric BMI was calculated using the conventional formula (weight in kilograms divided by height in meters squared). Child anthropometric BMI z-scores will be included as an additional dependent variable to contextualize and benchmark DNAm-derived BMI risk scores.

### Covariate Measures

Child’s race, ethnicity, and sex were collected via caregiver report. These variables were included as covariates to account for demographic factors that may be associated with differential exposure to social, environmental, and structural conditions linked to child health outcomes and DNAm-based measures.

### Statistical analysis plan

The current DNAm sub-study began after the initiation of the primary RCT, resulting in the current study being an observational secondary analysis of participants in the original iPCIT trial. As a result, the current study consisted of a larger proportion of missing data at baseline than at the 12-month follow-up. For this reason, we conducted analyses focused on this final follow-up for our primary outcome. In the current study, participants were included if they had DNAm data that met quality-control thresholds for at least one time point.

We conducted path analysis models in R (R Core Team, 2025) using the lavaan package (Rosseel, 2012). To address missing data, full-information maximum likelihood (FIML) estimation was used to retain all available data, and maximum likelihood estimation with robust standard errors was applied. Sensitivity analyses were conducted to determine whether conclusions were consistent across unadjusted observed-data analyses and FIML-estimated models. Our primary model included treatment condition (iPCIT or RAU) as the primary predictor, DNAm-derived BMI risk score at the 12-month follow-up as the primary outcome, and controlled for baseline DNAm-derived BMI, child sex, and child race and/or ethnicity, with all baseline variables allowed to correlate with one another. All path models included child sex, child race, and child ethnicity as covariates. Covariates were selected based on their known association with DNAm or their differences between the two treatment conditions.

We also examined the variation in treatment effects between DNAm-derived BMI and measured anthropometric BMI by conducting path analysis models that incorporated each outcome and its covariance within a single framework. These analyses were conducted using path models in which DNAm-derived BMI and anthropometric BMI at 12-months were specified as parallel outcome variables, each controlling for its respective baseline values.

In addition to examining direct effects of treatment on DNAm-derived BMI, we tested a mediation model in which changes in parenting behaviors (i.e., increases in responsive and consistent parenting from baseline to posttreatment and/or decreases in harsh and inconsistent parenting, assessed via DPICS-IV observations) function as mediators of treatment effects on BMI outcomes. All parenting behaviors were observed during child-directed play. Responsive and consistent parenting behaviors were assessed using “Do” skills, which include praise, behavior descriptions, and reflections; harsh and inconsistent parenting behaviors were assessed using “Don’t” skills, which include questions, commands, and criticisms. Mediation models examining the two parenting domains were estimated separately in R, with posttreatment “Do” and “Don’t” skills as predictors and controlling for their respective baseline measures. Indirect effects were estimated using bias-corrected bootstrapped confidence intervals, with 2000 bootstrap samples to ensure stable estimates. This approach allowed us to examine whether improvements in observed parenting accounted for reductions in DNAm-derived BMI risk scores among families receiving iPCIT.

## Results

### Participants

The final sample included 74 participants (*n* = 35 iPCIT; *n* = 39 RAU). At baseline, children were on average 36-months-old, with no age differences between conditions, and the sample was predominantly male (71.6%) across both groups. The majority of participants identified as Hispanic/Latino (71.1%), followed by Non-Hispanic Black/African American (18.9%). Primary caregivers were predominantly women (94.6%), and most primary caregivers reported English as their preferred language (60.8%), although a substantial proportion preferred Spanish (39.2%). Caregivers’ educational attainment was varied, with 43.2% reporting a college degree or higher. Participating families represented a range of socioeconomic backgrounds, with almost two-thirds of the sample (62.2%) classified as having below-adequate economic resources based on calculated income-to-needs ratios (calculated by dividing annual household income by the official poverty threshold for household size). At baseline, the majority of children fell within the healthy-weight range for age and sex, with relatively few children meeting criteria for overweight or obesity classifications. Thus, the present study focused on variation in DNAm-derived BMI risk scores as an early biological indicator of cardiometabolic risk rather than existing obesity status. Demographic characteristics were generally comparable across conditions, with some variation in racial/ethnic composition, language preference, and income-to-needs categorization. See **Table 1** for full demographic information.

### Primary Analyses

Primary analyses tested whether assignment to iPCIT was associated with lower DNAm-derived BMI scores at the 12-month follow-up, after accounting for baseline DNAm-derived BMI and selected covariates. See **Table 2** for complete results. For DNAm BMI at the 12-month follow-up, statistically significant treatment differences were observed after accounting for baseline DNAm-derived BMI. The iPCIT group (*M* = -0.53; *SD* = 0.10) demonstrated lower DNAm-derived BMI at 12 months relative to the RAU group (*M* = -0.49; *SD* = 0.10), *(b* = -0.06, 95% CI [-0.11, -0.01], *p* = 0.01), with a small-to-medium effect size (Cohen’s d = 0.42). Baseline DNAm-derived BMI was positively associated with 12-month DNAm-derived BMI group *(b* = 0.29, 95% CI [-0.02, 0.60], *p* = 0.07). Neither child sex nor child race and/or ethnicity was a significant predictor of DNAm BMI at 12 months (child sex: *b* = -0.02, 95% CI [-0.08, 0.03], *p* = 0.37; child race and/or ethnicity: *b* = 0.03, 95% CI [-0.07, 0.13], *p* = 0.56).

As a sensitivity analysis, we repeated the primary model adjusting for family income-to-needs ratio. Inclusion of income-to-needs did not alter the pattern of results. Treatment condition remained significantly associated with lower DNAm-derived BMI risk scores at the 12-month follow-up (*b* = -0.05, 95% CI [-0.10, -0.01], *p* = 0.03). Income-to-needs ratio was not independently associated with DNAm-derived BMI risk scores (*b* = 0.01, 95% CI [-0.01, 0.03], *p* = 0.30).

Parallel models were used to examine whether treatment-related differences were specific to DNAm-derived BMI or also evident with anthropometric BMI. These models included the additional variable of anthropometric BMI at 12 months (*M* = 16.12; *SD* = 2.04), accounting for baseline anthropometric BMI (*M* = 16.78; *SD* = 2.11). At 12-months, DNAm-derived BMI was not significantly associated with anthropometric BMI (*b* = -0.01, 95% CI [-0.01, 0.02], *p* = 0.58). However, baseline-anthropometric BMI, strongly predicted 12-month anthropometric BMI (*b* = 0.71, 95% CI [0.15, 1.27], *p* = 0.01) See **Figure 1**. treatment effect on DNAm-derived BMI remained significant after accounting for anthropometric BMI (*b* = -0.06, 95% CI [-0.11, -0.01], *p* = 0.02). In contrast, no treatment effect was observed for anthropometric BMI at 12-months (*b* = -0.10, 95% CI [-0.62, 0.43], *p* = 0.72). See **Table 3** for complete results.

### Mediation Analyses

We conducted two separate mediation models to examine whether post-treatment parenting behaviors mediated the treatment effect on DNAm-derived BMI at 12-months. One model included DPICS-IV “Do” behaviors (i.e., responsive and consistent parenting practices) as the mediator, and a second model included DPICS-IV “Don’t” behaviors (i.e., harsh and inconsistent parenting practices) as the mediator. Both models included their respective parenting behaviors (“Do”/ “Don’t”) at baseline as covariates. Results indicated that iPCIT was significantly associated with increases in “Do” behaviors (*b* = 0.49, 95% CI [0.03, 0.94], *p* = 0.04). However, “Do” behaviors did not significantly predict DNAm-derived BMI at 12-months (*b* = -0.02, 95% CI [-0.04, 0.01], *p* = 0.12), and the indirect effect was not significant (*b* = -0.01, 95% CI [-0.03, 0.00], *p* = 0.30). In the model testing “Don’t” behaviors as the mediator, iPCIT significantly predicted decreases in “Don’t” behaviors (*b* = -0.49, 95% CI [-0.92, -0.04], *p* = 0.03). However, “Don’t” behaviors did not significantly predict DNAm-derived BMI at 12 months (*b* = -0.01, 95% CI [-0.04, 0.02], *p* = 0.33), and the indirect effect was not significant (*b* = 0.01, 95% CI [-0.01, 0.02], *p* = 0.38).

## Discussion

The current study represents one of the first examinations of whether a parenting intervention influences DNAm-derived BMI risk scores in early childhood. Children who participated in the iPCIT condition demonstrated significantly lower DNAm-derived BMI risk scores at the 12-month follow-up compared to those in the control condition. This treatment effect remained consistent even after controlling for potential confounders such as child sex and race and/or ethnicity, suggesting that the observed treatment effect is robust and not driven by these demographic characteristics. Notably, the observed effects emerged from an intervention targeting the broader caregiving environment rather than weight-related behaviors specifically. This finding is consistent with the notion that responsive and consistent parenting and household structure may serve as foundational conditions that support healthy developmental and physiological processes, even when health behaviors are not explicitly targeted. Further, these findings align with a growing body of literature demonstrating that parenting and family-based interventions can influence children’s DNAm profiles across a range of health-related outcomes.^34,35,37,38^

Findings from the current study align with prior studies showing that similar parenting interventions are associated with changes in DNAm-based risk markers linked to stress-related biological aging.^34^ Additionally, previous findings with the same cohort demonstrated that iPCIT was associated with changes in DNAm-accelerated aging and inflammation.^35,37^ Further, the current study provides findings consistent with previous work examining the positive impact of general parenting interventions in mitigating child cardiometabolic risk.^32,33^ Importantly, this pattern extends prior work linking parenting practices to children’s weight status and cardiometabolic health,^25,26^ by suggesting a potential biological pathway through which these associations may operate. Consistent with the guiding bioecological framework, these findings support the idea that proximal caregiving environments can become biologically embedded, shaping early indicators of cardiometabolic risk. In this way, the present results build on existing observational and intervention research by demonstrating that parenting practices may shape obesity-related risk at the molecular level, even in early childhood.

In the current study, the observed treatment effect on DNAm-derived BMI risk did not extend to anthropometric BMI. Specifically, while the iPCIT condition was associated with lower DNAm-derived BMI risk scores, there was no corresponding effect on children’s anthropometric BMI at the 12-month follow-up. Moreover, DNAm-derived BMI and anthropometric BMI were not significantly correlated in this sample, suggesting that these indices may capture distinct aspects of cardiometabolic risk in early childhood.^56^ These findings are consistent with emerging evidence that DNAm-based biomarkers may capture early biological embedding of risk processes that are not yet observable at the phenotypic level, particularly in young children.^57^ One possibility is that DNA-based measures may provide a more sensitive indicator of early intervention effects, whereas changes in measured BMI may require longer developmental windows to become detectable.

The absence of a correlation between DNAm-derived BMI and anthropometric BMI may indicate limited construct validity of the DNAm score in salivary tissue among preschool-aged children, rather than heightened sensitivity to early biological risk processes. Prior work by Raffington et al.^24^ demonstrated modest associations between salivary DNAm-BMI scores, anthropometric BMI, and socioeconomic status in older youth. The extent to which these findings generalize to preschool-aged populations, however, remains unclear. Importantly, the McCartney DNAm-derived BMI risk score was originally developed using blood-based methylation data from primarily European ancestry adult samples, raising additional questions regarding its developmental, tissue-specific, and ancestral generalizability when applied to salivary DNAm data in a diverse sample of young children. Therefore, although the observed discrepancy may reflect epigenetic processes that emerge prior to phenotypic manifestations of obesity risk, this interpretation should be considered preliminary until additional validation studies are conducted in early childhood samples.

In the study that developed the current DNAm-derived BMI risk score,^55^ there was only a modest association with anthropometric BMI (explaining 12.5% of variance in BMI), suggesting that, even in adult samples, the DNAm-derived risk score may be tapping into molecular processes that reflect biological risk for future weight gain or metabolic dysregulation rather than current body size. Further, the DNAm-derived risk score used in the current study was not developed based on childhood metabolic risk; instead, the CpG sites and their regression weights reflect the methylation architecture of adiposity in mature metabolic systems. Future research would benefit from using longitudinal pediatric cohorts with repeated methylation and anthropometric assessments to build DNAm-derived risk scores that reflect how cardiometabolic health develops across childhood, rather than assuming that DNAm-derived BMI risk measurements remain consistent from childhood to adulthood. Further, future research that tracks DNAm-derived BMI across developmental stages, particularly across pubertal development and adolescence, would help us better understand if DNAm-derived BMI prospectively predicts future metabolic risk.

Notably, while iPCIT was significantly associated with improvements in responsive and consistent parenting practices and with reductions in harsh and inconsistent parenting practices, as measured by the DPICS-IV “Do” and “Don’t” behaviors, these parenting behaviors did not mediate the relationship between treatment condition and DNAm-derived BMI risk at 12 months. Although the iPCIT produced measurable changes in observed parenting behaviors, these changes do not appear to directly account for the observed reductions in DNAm-based cardiometabolic risk. These findings are inconsistent with prior research demonstrating that changes in specific parenting behaviors (e.g., shouting, commands, praise) directly affect children’s DNAm-derived risk scores.^37,38^ However, these null mediation findings should be interpreted cautiously given the relatively small sample size (*n* = 74), which may have limited statistical power for the association between post-treatment observed parenting and epigenetic BMI at follow-up. Importantly, the observed associations were generally in the expected direction, suggesting that the absence of statistically significant mediation may reflect limited power for these higher-order analyses, rather than a true absence of parenting-related biological effects. Further, previous findings from this sample demonstrated associations between iPCIT participation and improvements in DNAm-derived inflammation and epigenetic aging,^35,37^ supporting stress regulation as a plausible mechanism linking caregiving quality to DNAm-based cardiometabolic risk. However, the DPICS-IV primarily indexes intervention-specific parenting skills and treatment fidelity and may therefore be less sensitive to broader, sustained aspects of the caregiving environment that are more likely to influence biological embedding. Future studies using repeated assessments and measures that more accurately capture broadband parenting practices and family processes may provide greater insight into the mechanisms by which caregiving experiences shape epigenetic risk profiles.

Several alternative mechanisms may help explain the lack of significant mediation found for parenting practices. First, the intervention may have influenced children’s cardiometabolic risk through direct changes in physiological stress regulation. Improvements in the caregiving environment could reduce chronic activation of stress-response systems (e.g., HPA axis activity),^58^ which in turn may shape DNAm patterns associated with metabolic risk^59^. These physiological processes may operate somewhat independently of the specific parenting behaviors captured in the DPICS-IV observational codes, given that these parenting behaviors are specific to the intervention (e.g., specific praise, active ignoring) rather than broadband parenting domains (e.g., warmth, supportiveness, behavioral control). Additionally, changes in family routines and specific health-related behaviors may represent an additional pathway. Parenting interventions such as iPCIT may indirectly influence children’s sleep patterns, dietary habits, and physical activity through increased structure and consistency.^60,61^ These factors are well-established contributors to cardiometabolic health and may be more directly linked to DNAm-derived BMI risk than the broader parenting domains assessed in our coding (e.g., praise, reflection, criticism), which do not specifically target health-related parenting behaviors. Finally, it is also possible that unmeasured caregiver-level changes, such as reductions in parental stress, depression, or family discordance, played a role in shaping children’s overarching biological risk.^28,38,62^ Improvements in caregiver well-being and the broader home environment may create conditions that support healthier physiological functioning in children and reduce cardiometabolic risk,^21,32,63^ even if these changes are not fully captured by observed parenting behaviors during structured interactions.

## Strengths, Limitations & Future Directions

Current study strengths include the longitudinal randomized controlled trial design within an understudied sample exposed to elevated stress during early childhood. Data were collected at multiple levels of analysis (e.g., demographic, observational, and biological), enabling a more comprehensive assessment of child and family functioning. A key strength of the present study is the use of an innovative biomarker of cardiometabolic risk rather than relying solely on traditional anthropometric indices. DNAm BMI in the current study was derived from saliva samples, representing a non-invasive and developmentally appropriate method for assessing biological risk in pediatric populations. This approach enhances participant acceptability, especially in high-risk or underserved populations where more invasive procedures may present barriers to participation. Looking forward, these non-invasive epigenetic markers may hold promise for early risk monitoring, identifying children who could benefit from targeted prevention efforts, and evaluating intervention-related changes in biological risk over time.

Finally, the families who participated in this study represent large and growing populations within the U.S., including Latino families who remain underrepresented in epigenomics research and children with developmental delays, who remain underrepresented in prevention and intervention studies. Firstly, the lack of Latino representation in epigenomics limits understanding of how social and environmental exposures shape biological processes across diverse populations and may contribute to inequities in risk prediction, intervention effectiveness, and, most importantly, protective factors for health status.^41^ Focusing on these groups is especially important given their disproportionate exposure to structural inequities, including socioeconomic disadvantage and neighborhood environments, which contribute to elevated risk for poor cardiometabolic health outcomes.^4,64,65^ Second, children with developmental delays are often underrepresented in prevention and intervention research and experience persistent disparities in identification and access to care^66–68^ despite evidence that early family-based interventions can meaningfully influence developmental and biological outcomes,^42^ highlighting the importance of including these populations in studies of risk factors that are responsive to early intervention.

The present study has several limitations that should be considered when interpreting the findings. First, analyses were conducted on a subsample of the larger randomized controlled trial. DNAm data were collected from only 74 of the 150 families enrolled in the parent study, as the epigenetic substudy was initiated after the parent RCT had begun. Prior published work on this subsample suggests that participants included in the biomarker subsample did not differ significantly from those in the full trial on demographic characteristics or key outcomes, thereby supporting the representativeness of the sample relative to the full set of participants in the RCT.^35^ The reduced sample size nonetheless raises concern regarding statistical power. As a result, the study may have been underpowered to detect smaller effects, including indirect or mechanistic pathways (e.g., mediation). Additionally, while child sex was equally distributed across treatment conditions (iPCIT and RAU), boys comprised nearly three-quarters of the sample. Considering that boys are more likely to be referred for behavioral concerns,^69^ this distribution is not unexpected. However, this may limit the generalizability of our findings across sexes. Further, parenting data largely reflected maternal behaviors. This limitation is common in developmental and clinical psychology research, underscoring the need for greater attention to other caregivers (e.g., fathers^70,71^) in future work.

Finally, the present study did not include measures of health-related behaviors that are proximal indicators of cardiometabolic risk, such as dietary patterns and physical activity. Although the present study included caregiver-reported child sleep measures, these measures were relatively limited in scope and may not have fully captured children’s sleep patterns.^72^ Therefore, our ability to identify the behavioral pathways through which these effects may have occurred remains limited. In addition, the study did not assess parenting practices specific to these domains (e.g., feeding-related parenting, support for physical activity, nighttime parenting), which may represent key mechanisms linking intervention effects to child health behaviors and downstream biological risk. Future research incorporating multi-domain assessments of child health behaviors alongside domain-specific parenting practices will be important for clarifying these pathways. Such work may also benefit from qualitative approaches to better understand family beliefs and practices around health behaviors, informing more targeted prevention and intervention strategies for youth cardiometabolic health.

## Conclusions

Altogether, this study provides preliminary evidence that early parenting interventions focused on improving parent-child interactions can influence children’s DNAm markers related to cardiometabolic risk, even in the absence of immediate changes in measured BMI. These results highlight the potential for epigenetic biomarkers to serve as early, sensitive indicators of intervention effects. Future research should build on these findings by employing a more comprehensive, multi-level approach that integrates biological, behavioral, and psychosocial data. Future research may also incorporate focus groups and participatory action methods to engage underserved families directly, ensuring that family-centered interventions align with their cultural values, lived experiences, and specific needs. In sum, understanding how family dynamics and parenting practices specifically shape biological risk, as reflected by epigenetic BMI, will be critical for designing culturally responsive, family-centered prevention strategies.

## Data Availability

The datasets generated and/or analyzed during the current study are available from the corresponding author on reasonable request.

## Author contributions

Adamari Lopez (Conceptualization, Writing—original draft, Writing—review & editing);

Sarah M. Merrill (Data curation, Writing—review & editing);

Anne K. Bozack (Data curation, Writing—review & editing);

Andres Cardenas (Data curation, Writing—review & editing);

Jonathan S. Comer (Conceptualization, Data curation, Funding acquisition, Investigation, Methodology, Project administration, Resources, Writing – review & editing);

Daniel M. Bagner (Conceptualization; Data curation; Funding acquisition; Investigation; Methodology; Project administration; Resources; Writing – review & editing);

April Highlander (Writing—review & editing);

Justin Parent (Conceptualization; Data curation; Formal analysis; Funding acquisition; Investigation; Methodology; Project administration; Resources; Supervision; Writing – review & editing)

## Ethics Approval

This study was approved by the Florida International University Institutional Review Board, and informed, written consent was obtained from all participants.

## References

1. Skinner, A. C., Ravanbakht, S. N., Skelton, J. A., Perrin, E. M. & Armstrong, S. C. Prevalence of Obesity and Severe Obesity in US Children, 1999–2016. Pediatrics 141, e20173459 (2018).

2. Stierman, B. et al. Changes in adiposity among children and adolescents in the United States, 1999–2006 to 2011–2018. Am. J. Clin. Nutr. 114, 1495–1504 (2021).

3. Cunningham, S. A. et al. Changes in the Incidence of Childhood Obesity. Pediatrics 150, e2021053708 (2022).

4. Vazquez, C. E. & Cubbin, C. Socioeconomic Status and Childhood Obesity: a Review of Literature from the Past Decade to Inform Intervention Research. Curr. Obes. Rep. 9, 562–570 (2020).

5. Chung, Y. L. & Rhie, Y.-J. Severe Obesity in Children and Adolescents: Metabolic Effects, Assessment, and Treatment. J. Obes. Metab. Syndr. 30, 326–335 (2021).

6. Isasi, C. R., Rastogi, D. & Molina, K. Health issues in Hispanic/Latino youth. J. Lat. Psychol. 4, 67–82 (2016).

7. Singh, A. S., Mulder, C., Twisk, J. W. R., Van Mechelen, W. & Chinapaw, M. J. M. Tracking of childhood overweight into adulthood: a systematic review of the literature. Obes. Rev. 9, 474–488 (2008).

8. Simmonds, M., Llewellyn, A., Owen, C. G. & Woolacott, N. Predicting adult obesity from childhood obesity: a systematic review and meta-analysis. Obes. Rev. 17, 95–107 (2016).

9. Biro, F. M. & Wien, M. Childhood obesity and adult morbidities. Am. J. Clin. Nutr. 91, 1499S–1505S (2010).

10. Quek, Y., Tam, W. W. S., Zhang, M. W. B. & Ho, R. C. M. Exploring the association between childhood and adolescent obesity and depression: a meta-analysis. Obes. Rev. 18, 742–754 (2017).

11. Rankin, J. et al. Psychological consequences of childhood obesity: psychiatric comorbidity and prevention. Adolesc. Health Med. Ther. Volume 7, 125–146 (2016).

12. Wu, Y., Li, D. & Vermund, S. H. Advantages and Limitations of the Body Mass Index (BMI) to Assess Adult Obesity. Int. J. Environ. Res. Public. Health 21, 757 (2024).

13. Rothman, K. J. BMI-related errors in the measurement of obesity. Int. J. Obes. 32, S56–S59 (2008).

14. Nimptsch, K., Konigorski, S. & Pischon, T. Diagnosis of obesity and use of obesity biomarkers in science and clinical medicine. Metabolism 92, 61–70 (2019).

15. Rushing, A., Sommer, E. C., Zhao, S., Po’e, E. K. & Barkin, S. L. Salivary epigenetic biomarkers as predictors of emerging childhood obesity. BMC Med. Genet. 21, 34 (2020).

16. Boyce, W. T. & Kobor, M. S. Development and the epigenome: the ‘synapse’ of gene–environment interplay. Dev. Sci. 18, 1–23 (2015).

17. Bronfenbrenner, U. & Ceci, S. J. Nature-nuture reconceptualized in developmental perspective: A bioecological model. Psychol. Rev. 101, 568–586 (1994).

18. Gallo, L. C. et al. Social, psychological, and cultural dimensions of cardiovascular health among Hispanic/Latino adults: A narrative review of findings from the Hispanic Community Health Study/Study of Latinos. Health Psychol. 10.1037/hea0001562(2025) doi:10.1037/hea0001562.

19. Campbell, M. K. Biological, environmental, and social influences on childhood obesity. Pediatr. Res. 79, 205–211 (2016).

20. Jebeile, H., Kelly, A. S., O’Malley, G. & Baur, L. A. Obesity in children and adolescents: epidemiology, causes, assessment, and management. Lancet Diabetes Endocrinol. 10, 351–365 (2022).

21. Halliday, J. A., Palma, C. L., Mellor, D., Green, J. & Renzaho, A. M. N. The relationship between family functioning and child and adolescent overweight and obesity: a systematic review. Int. J. Obes. 38, 480–493 (2014).

22. Kaufman, J. et al. Adverse Childhood Experiences, Epigenetic Measures, and Obesity in Youth. J. Pediatr. 202, 150–156.e3 (2018).

23. Reed, Z. E., Suderman, M. J., Relton, C. L., Davis, O. S. P. & Hemani, G. The association of DNA methylation with body mass index: distinguishing between predictors and biomarkers. Clin. Epigenetics 12, 50 (2020).

24. Raffington, L. et al. Salivary Epigenetic Measures of Body Mass Index and Social Determinants of Health Across Childhood and Adolescence. JAMA Pediatr. 177, 1047 (2023).

25. Melis Yavuz, H. & Selcuk, B. Predictors of obesity and overweight in preschoolers: The role of parenting styles and feeding practices. Appetite 120, 491–499 (2018).

26. Pinquart, M. Associations of General Parenting and Parent–Child Relationship With Pediatric Obesity: A Meta-Analysis. J. Pediatr. Psychol. 39, 381–393 (2014).

27. Kiefner-Burmeister, A. & Hinman, N. The Role of General Parenting Style in Child Diet and Obesity Risk. Curr. Nutr. Rep. 9, 14–30 (2020).

28. Hogan, C. M. et al. The Impact of Early Life Adversity on Peripubertal Accelerated Epigenetic Aging and Psychopathology. J. Am. Acad. Child Adolesc. Psychiatry 64, 724–733 (2025).

29. Parent, J., McKee, L. G. N. Rough, J. & Forehand, R. The Association of Parent Mindfulness with Parenting and Youth Psychopathology Across Three Developmental Stages. J. Abnorm. Child Psychol. 44, 191–202 (2016).

30. Metrailer, G. et al. Community threat, positive parenting, and accelerated epigenetic aging: Longitudinal links from childhood to adolescence. Child Dev. 97, 9–20 (2026).

31. Domoff, S. E. & Niec, L. N. Parent-child interaction therapy as a prevention model for childhood obesity: A novel application for high-risk families. Child. Youth Serv. Rev. 91, 77–84 (2018).

32. White, H. I., Holmbeck, K., Ratmansky, J., Kong, K. L. & Anzman-Frasca, S. A Systematic Review of Early General Parenting Interventions: Long-term Effects in Underrepresented Populations and Implications for Obesity Prevention. Curr. Obes. Rep. 13, 789–816 (2024).

33. Kong, K. L. et al. Systematic Review of General Parenting Intervention Impacts on Child Weight as a Secondary Outcome. Child. Obes. 19, 293–308 (2023).

34. Creasey, N., Leijten, P., Overbeek, G. & Tollenaar, M. S. Incredible years parenting program buffers prospective association between parent-reported harsh parenting and epigenetic age deceleration in children with externalizing behavior. Psychoneuroendocrinology 165, 107043 (2024).

35. Merrill, S. M. et al. Telehealth Parenting Program and Salivary Epigenetic Biomarkers in Preschool Children With Developmental Delay: NIMHD Social Epigenomics Program. JAMA Netw. Open 7, e2424815 (2024).

36. Sullivan, A. D. W. et al. Intervening After Trauma: Child–Parent Psychotherapy Treatment Is Associated With Lower Pediatric Epigenetic Age Acceleration. Psychol. Sci. 35, 1062–1073 (2024).

37. Sullivan, A. D. W. et al. Parenting Practices May Buffer the Impact of Adversity on Epigenetic Age Acceleration Among Young Children With Developmental Delays. Psychol. Sci. 34, 1173–1185 (2023).

38. Brody, G. H., Yu, T., Chen, E., Beach, S. R. H. & Miller, G. E. Family-centered prevention ameliorates the longitudinal association between risky family processes and epigenetic aging. J. Child Psychol. Psychiatry 57, 566–574 (2016).

39. Zheng, M. et al. Association Between Longitudinal Trajectories of Lifestyle Pattern and BMI in Early Childhood. Obesity 29, 879–887 (2021).

40. Guerrero, A. D. et al. Racial and Ethnic Disparities in Early Childhood Obesity: Growth Trajectories in Body Mass Index. J. Racial Ethn. Health Disparities 3, 129–137 (2016).

41. Gillman, A. S., Pérez-Stable, E. J. & Das, R. Advancing Health Disparities Science Through Social Epigenomics Research. *JAMA Netw*. Open 7, e2428992 (2024).

42. Bagner, D. M. et al. Telehealth Treatment of Behavior Problems in Young Children With Developmental Delay: A Randomized Clinical Trial. JAMA Pediatr. 177, 231 (2023).

43. Comer, J. S. et al. Remotely delivering real-time parent training to the home: An initial randomized trial of Internet-delivered parent–child interaction therapy (I-PCIT). J. Consult. Clin. Psychol. 85, 909–917 (2017).

44. Grondhuis, S. N. & Aman, M. G. Overweight and obesity in youth with developmental disabilities: a call to action. J. Intellect. Disabil. Res. 58, 787–799 (2014).

45. Emerson, E. & Brigham, P. Exposure of children with developmental delay to social determinants of poor health: cross-sectional case record review study. Child Care Health Dev. 41, 249–257 (2015).

46. Achenbach, T. M. Manual for the Child Behavior Checklist/4-18 and 1991 profile. Univ. Vt. Dep. Psychiatry (1991).

47. Constantino, J. N. Social Responsiveness Scale. in Encyclopedia of Autism Spectrum Disorders (ed. Volkmar, F. R.) 4457–4467 (Springer International Publishing, Cham, 2021). doi:10.1007/978-3-319-91280-6_296.

48. Wechsler, D. Wechsler Abbreviated Scale of Intelligence--Second Edition. 10.1037/t15171-000 (2018).

49. Lieneman, C., Brabson, L., Highlander, A., Wallace, N. & McNeil, C. Parent–Child Interaction Therapy: current perspectives. Psychol. Res. Behav. Manag. Volume 10, 239–256 (2017).

50. Eyberg, S. M. Dyadic Parent-Child Interaction Coding System (DPICS): Comprehensive Manual for Research and Training. (PCIT International, Incorporated, 2013).

51. Aryee, M. J. et al. Minfi: a flexible and comprehensive Bioconductor package for the analysis of Infinium DNA methylation microarrays. Bioinformatics 30, 1363–1369 (2014).

52. Murat, K. et al. Ewastools: Infinium Human Methylation BeadChip pipeline for population epigenetics integrated into Galaxy. GigaScience 9, giaa049 (2020).

53. Fortin, J.-P. et al. Functional normalization of 450k methylation array data improves replication in large cancer studies. Genome Biol. 15, 503 (2014).

54. Zheng, S. C. et al. EpiDISH web server: Epigenetic Dissection of Intra-Sample-Heterogeneity with online GUI. Bioinformatics 36, 1950–1951 (2019).

55. McCartney, D. L. et al. Epigenetic prediction of complex traits and death. Genome Biol. 19, 136 (2018).

56. Hamilton, O. K. L. et al. An epigenetic score for BMI based on DNA methylation correlates with poor physical health and major disease in the Lothian Birth Cohort. Int. J. Obes. 43, 1795–1802 (2019).

57. Belsky, D. W. et al. DunedinPACE, a DNA methylation biomarker of the pace of aging. eLife 11, e73420 (2022).

58. Blair, C. et al. Allostasis and allostatic load in the context of poverty in early childhood. Dev. Psychopathol. 23, 845–857 (2011).

59. Bose, M., Oliván, B. & Laferrère, B. Stress and obesity: the role of the hypothalamic–pituitary–adrenal axis in metabolic disease. Curr. Opin. Endocrinol. Diabetes Obes. 16, 340–346 (2009).

60. Acosta, J., Garcia, D. & Bagner, D. M. Parent-Child Interaction Therapy for Children with Developmental Delay: The Role of Sleep Problems. J. Dev. Behav. Pediatr. 40, 183–191 (2019).

61. Chen, Y., Haines, J., Charlton, B. M. & VanderWeele, T. J. Positive parenting improves multiple aspects of health and well-being in young adulthood. Nat. Hum. Behav. 3, 684–691 (2019).

62. Natsuaki, M. N. et al. Raised by Depressed Parents: Is it an Environmental Risk? Clin. Child Fam. Psychol. Rev. 17, 357–367 (2014).

63. Ochoa, A. & Berge, J. M. Home Environmental Influences on Childhood Obesity in the Latino Population: A Decade Review of Literature. J. Immigr. Minor. Health 19, 430–447 (2017).

64. Chung, S. T., Krenek, A. & Magge, S. N. Childhood Obesity and Cardiovascular Disease Risk. Curr. Atheroscler. Rep. 25, 405–415 (2023).

65. Larson, N. I., Wall, M. M., Story, M. T. & Neumark-Sztainer, D. R. Home/family, peer, school, and neighborhood correlates of obesity in adolescents. Obesity 21, 1858–1869 (2013).

66. Guerrero, A. D., Rodriguez, M. A. & Flores, G. Disparities in Provider Elicitation of Parents’ Developmental Concerns for US Children. Pediatrics 128, 901–909 (2011).

67. Coker, T. R., Shaikh, Y. & Chung, P. J. Parent-Reported Quality of Preventive Care for Children At-Risk for Developmental Delay. Acad. Pediatr. 12, 384–390 (2012).

68. Rosenberg, S. A., Zhang, D. & Robinson, C. C. Prevalence of Developmental Delays and Participation in Early Intervention Services for Young Children. Pediatrics 121, e1503–e1509 (2008).

69. Coles, E. K., Slavec, J., Bernstein, M. & Baroni, E. Exploring the Gender Gap in Referrals for Children With ADHD and Other Disruptive Behavior Disorders. J. Atten. Disord. 16, 101–108 (2012).

70. Parent, J., Forehand, R., Pomerantz, H., Peisch, V. & Seehuus, M. Father Participation in Child Psychopathology Research. J. Abnorm. Child Psychol. 45, 1259–1270 (2017).

71. Yaremych, H. E. & Persky, S. Recruiting Fathers for Parenting Research: An Evaluation of Eight Recruitment Methods and an Exploration of Fathers’ Motivations for Participation. Parenting 23, 1–32 (2023).

72. Warner, M., Gillenson, C. J., Parent, J., Comer, J. S. & Bagner, D. M. Internet-delivered Parent-Child Interaction Therapy and Sleep Quality in Children With Developmental Delay: Examining the Mediating Role of Bedtime Resistance Behaviors. J. Dev. Behav. Pediatr. 46, e269–e274 (2025).

