## Supplementary material for "The Role of Parenting in Mitigating Epigenetic Cardiometabolic Risk in a Sample of Predominantly Latino Preschoolers": Tables & Figures

**Table 1**
*Sociodemographic Characteristics of Participants at Baseline by Treatment Condition.*
iPCIT = internet-based Parent-Child Interaction Therapy; RAU = referrals as usual.

| | iPCIT (*n* = 35) | | RAU (*n* = 39) | | Total (*N* = 74) | |
|---|---|---|---|---|---|---|
| | *M* | SD | *M* | *SD* | *M* | *SD* |
| Child Age | 36.1(0.70) | | 36.1(0.67) | | 36.1(0.68) | |
| Child Baseline BMI | 16.3(1.52) | | 16(2.42) | | 16.12(2.04) | |
| Child DNAm-derived BMI | -0.43(0.12) | | -0.49(0.11) | | -0.46(0.11) | |
| | *n* | % | *n* | % | *n* | % |
| Child Sex | | | | | | |
| Male | 25(71.4) | | 28(71.8) | | 53(71.6) | |
| Female | 10(28.6) | | 11(28.2) | | 21(28.4) | |
| Child ethnicity and race | | | | | | |
| Non-Hispanic Asian | 1(2.8) | | 2(5.1) | | 3(4.1) | |
| Non-Hispanic Black/African American | 4(11.4) | | 10(25.6) | | 14(18.9) | |
| Black Latino or Hispanic | 1(2.8) | | 2(5.1) | | 3(4.1) | |
| White Latino or Hispanic | 28(80.0) | | 22(56.4) | | 50(67.6) | |
| Non-Hispanic White | 1(2.8) | | 3(7.7) | | 4(5.4) | |
| Primary caregiver gender | | | | | | |
| Women | 33(94.3) | | 37(94.9) | | 70(94.6) | |
| Men | 2(5.7) | | 2(5.1) | | 4(5.4) | |
| Primary caregiver preferred language | | | | | | |
| English | 18(51.4) | | 27(69.2) | | 45(60.8) | |
| Spanish | 17(48.6) | | 12(30.8) | | 29(39.2) | |
| Primary caregiver educational attainment | | | | | | |
| Less than high school | 4(11.4) | | 4(10.3) | | 8(10.8) | |

| | | | |
|---|---|---|---|
| High school graduate / GED | 8(22.9) | 1(2.6) | 9(12.2) |
| Some college | 9(25.7) | 16(41.0) | 25(33.8) |
| College graduate | 9(25.7) | 17(43.6) | 26(35.1) |
| Graduate degree | 5(14.3) | 1(2.6) | 6(8.1) |
| Household income-to-needs | | | |
| Extreme poverty | 1(3.0) | 2(5.3) | 3(4.1) |
| Poor | 13(39.4) | 7(18.4) | 21(28.4) |
| Low Income | 12(36.4) | 10(26.3) | 22(29.7) |
| Adequate | 5(15.2) | 14(36.8) | 19(25.7) |
| Affluent | 2(6.0) | 5(13.2) | 7(9.5) |
| Primary caregiver marital status | | | |
| Married | 20(57.1) | 27(69.2) | 47(63.5) |
| Separated | 2(5.7) | 1(2.6) | 3(4.1) |
| Divorced | - | 1(2.6) | 1(1.4) |
| Single, never married | 13(37.1) | 9(23.1) | 22(29.7) |
| Domestic Partnership | - | 1(2.6) | 1(1.4) |

*Note.* Participants were on average 36 months old, and participant age did not differ by condition. Income-to-needs was calculated based on the primary caregiver's report of household income.

**Table 2**
*Path Analysis Results for the Effect of Treatment on DNAm BMI*

| Models and Outcomes | *b* | *SE* | 95% CI | | *p* |
|---|---|---|---|---|---|
| | | | *LL* | *UL* | |
| DNAm BMI at 12 months | | | | | |
| Treatment [c] | -0.06 | 0.03 | -0.11 | -0.01 | 0.01 |
| Baseline DNAm BMI | 0.29 | 0.16 | -0.02 | 0.60 | 0.07 |
| Child sex[a] | -0.02 | 0.03 | -0.08 | 0.03 | 0.37 |
| Child race and/or ethnicity[b] | 0.03 | 0.05 | -0.07 | 0.13 | 0.56 |

*Note*. CI = confidence interval; *LL* = lower limit; *UL* = upper limit.
[a] Child sex was coded as 0 = male, 1 = female. [b] Race was coded for analyses as a binary variable, Non-Hispanic White, with 0 = Non-Hispanic White and 1 = all other racial/ethnic identities. [c] Treatment was coded as 0 = referrals as usual and 1 = internet-based parent-child interaction therapy.

**Table 3**

*Path Analysis Results for the Effect of Treatment on DNAm BMI and Measured BMI*

| Models and Outcomes | *b* | *SE* | 95% CI *LL* | 95% CI *UL* | *p* |
|---|---|---|---|---|---|
| Measured BMI at 12 months | | | | | |
| Treatment[c] | -0.10 | 0.27 | -0.62 | 0.43 | 0.72 |
| Baseline measured BMI | 0.71 | 0.29 | 0.15 | 1.27 | 0.01 |
| Child sex[a] | 0.46 | 0.29 | -0.12 | 1.03 | 0.12 |
| Child race and/or ethnicity[b] | -0.01 | 0.33 | -0.65 | 0.63 | 0.98 |
| DNAm BMI at 12 months | | | | | |
| Treatment | -0.06 | 0.03 | -0.11 | -0.01 | 0.02 |
| Baseline DNAm BMI | 0.29 | 0.16 | -0.03 | 0.60 | 0.08 |
| Child sex | -0.02 | 0.03 | -0.08 | 0.03 | 0.37 |
| Child race and/or ethnicity | 0.03 | 0.05 | -0.07 | 0.13 | 0.52 |

*Note*. CI = confidence interval; *LL* = lower limit; *UL* = upper limit. BMI was calculated using the standardized formula: BMI = weight (lb) / height (in)$^2$ × 703.

[a] Child sex was coded as 0 = male, 1 = female. [b] Race was coded for analyses as a binary variable, Non-White Latino, with 0 = Non-Hispanic White and 1 = all other racial/ethnic identities. [c] Treatment was coded as 0 = referrals as usual and 1 = internet-based parent-child interaction therapy.

**Figure 1**

*Baseline and 12-month BMI variable correlations*

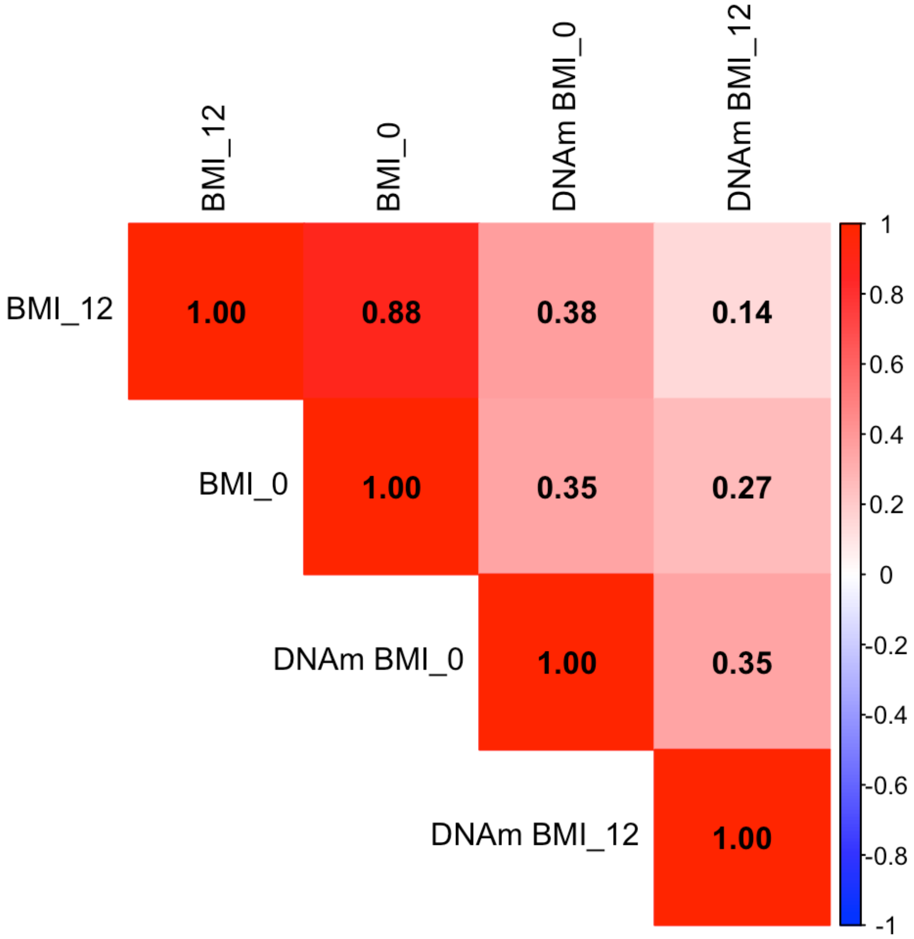


*Note.* BMI_0 = Baseline anthropometric BMI. BMI_12 = 12-month anthropometric BMI. DNAm BMI_0 = Baseline DNAm-derived BMI. DNAm BMI_12 = 12-month DNAm-derived BMI.
